# Institutional Trust in Times of Corona

**DOI:** 10.1101/2022.07.28.22278140

**Authors:** Erik Snel, Btissame El Farisi, Godfried Engbersen, André Krouwel

## Abstract

During the corona pandemic, governments of all countries appealed strongly to the trust of their populations by implementing drastic social and economic measures to prevent the spread of the virus. This study seeks to understand mechanisms that influence the level of institutional trust at the time of the corona pandemic. We are specifically interested in how three explanatory factors (socioeconomic status, experienced economic insecurity and dissatisfaction with the implemented corona policies) can, in mutual association, explain differences in institutional trust. This study is based on data from a large-scale panel survey on the social impact of COVID-19, carried out by Kieskompas research agency (N=22,696). Using a serial mediation analysis, we show that SES has both a direct and indirect effect on the level of institutional trust. People with higher SES experience less economic insecurity and have less dissatisfaction with the corona policies and, partly as a result of this, stronger institutional trust. It is also true that economic insecurity increases dissatisfaction with the corona policies and, partly as a result of this, weakens the level of trust.

## Introduction

During the corona pandemic of 2020, governments of all countries appealed strongly to the trust of their populations. Across the world, drastic measures were taken to reduce the number of COVID-19 infections and to prevent hospitals and other care organisations from becoming overburdened.Measures ranged from cancelling festivals and large sports events, to shutting down the hospitality sector, shops, schools and universities, up to imposing a night-time curfew. For the Dutch population it was the first time since the Second World War that they were forbidden from leaving their house at night. Although public disorder incidents occurred in several countries, including the Netherlands (the ‘curfew riots’ of late January 2020 stand out), overall the populations were compliant. One important reason pertains to the surge in trust in political institutions during the crisis, both in the Netherlands and in other affected countries [1-4].

This sudden and sometimes substantial increase in political trust in times of crisis is known as the ‘rally around the flag’ effect: in the event of external threats such as war, terrorist attacks or natural disasters, people collectively throw their weight behind their political leaders and institutions [5-8]. Rally effects are only temporary, however. Also with respect to the ‘corona-rally’, political trust initially increased strongly after the virus outbreak in the spring of 2020, to subsequently reduce again [9]. This also emerges from our research. In April 2020, 69 per cent of our respondents had (strong) trust in the national government and 60 per cent in their local government. These numbers had dropped to 55 and 50 per cent, respectively, in November 2020, and dropped further to a meagre 49 and 45 per cent, respectively, in March 2021 [10: 20].

Although this paper is not about fluctuations in political trust, the decreasing trust in the government and in important public health bodies such as the RIVM (Dutch National Institute for Public Health and the Environment) and GGD (municipal health service) can become problematic, especially in a long-term crisis such as the corona pandemic. One reason for this steadily decreasing political trust is thought to be the increasing doubts about and criticism of the corona policies pursued by the Dutch government [1]. After all, trust is situational and often strongly related to perceived procedural justice. Other factors also contribute to decreased institutional trust in times of corona. Research showed, for instance, that the consequences of the pandemic are unequally divided in society [10, 11]. That is why factors such as socioeconomic status and economic insecurity could also affect the level of institutional trust.

This paper seeks to understand the factors that jointly can explain the level of trust: The socioeconomic status of people, the extent to which the pandemic creates economic insecurity, and the subjective evaluation of, or discontent with, the government’s corona policies. This brings us to the following two-pronged problem definition of this study:

1. To what extent does institutional trust during the pandemic correlate with the respondents’ socioeconomic status?
2. To what extent is institutional trust during the pandemic influenced by specific experiences relating to the pandemic (fear of income loss due to COVID-19) and to discontent with the government’s measures?

The data used in this article derive from a large-scale survey (N= 22,696) on the societal impact of COVID-19, conducted in the Netherlands in November 2020. The survey contained many questions about the pandemic’s possible social consequences, including the level of trust in public authorities and in important public health bodies such as the RIVM and GGD.

## Socioeconomic status and institutional trust in times of corona

### Political versus institutional trust

Institutional trust is the main dependent variable in this study. The literature often distinguishes between political and institutional trust. Political trust pertains to people’s trust in political parties, in parliament or at the local level: the mayor and alderpersons and the municipal council. Institutional trust, on the other hand, pertains to trust in government institutions [cf. 12, 13]. In the survey, we asked about the trust that citizens have in the national and local government *and* in important public health bodies (RIVM and GGD). Another dimension of trust is interpersonal trust or trust in people in general (general trust). This will not be discussed in this article.

### Social status and institutional trust

It is a classic sociological premise that political or institutional trust correlates with social characteristics, in particular with education level, income and social class, labour market position (employed or unemployed), but also with health and the family situation (cohabiting or separated, with or without children). Research has shown that persons with a higher education level and higher income tend to have more political and institutional trust, with the education level being a particularly important determinant. As Uslaner [14: 108] observed: “virtually every study of generalized trust, in every setting, has found that education is a powerful predictor of trust” [cf. 15-21]. A general satisfaction regarding one’s own economic situation also correlates positively with a stronger level of trust in political institutions and authorities [22,23]. Institutional distrust occurs more often among people in structurally vulnerable positions with little opportunities for upward social mobility. Negative experiences such as unemployment, perceived discrimination or poor health also have a negative impact on social and political trust and tend to consolidate the distrust [24-26]. More generally, Zmerli and Newton [26: 70] find that especially the so-called ‘winners’ stand out for a strong level of social trust (in other people) and in political and institutional trust: “…those in dominant majority groups, people of high class, status, income and education, the happy and satisfied, and individuals who benefit from better health and post-materialist security.”

The literature reports various reasons and mechanisms to explain the connection between social status and trust. People with a higher socioeconomic position generally tend to be more optimistic about their life and also more positive about others. People in a more vulnerable socioeconomic position, on the other hand, are more inclined to be cynical, pessimistic and distrustful, both towards others *and* the government [28]. The latter are additionally more prone to a poorer health status, less economic security and greater economic risks, have less means to protect themselves against these risks and feel less protected by public bodies [20]. People in higher socioeconomic positions instead have more reasons to trust the government and other institutions. They are better equipped to utilise political and social institutions based on their understanding of how they operate, and if necessary, they have more political self-confidence and opportunities to positively influence those institutions [27: 71]. The social resilience displayed by people in higher status groups results in greater interpersonal and institutional trust, compared to people in lower status groups. Or, as Putnam [18: 188] says: “In virtually all societies, ‘have-nots’ are less trusting than ‘haves’, probably because haves are treated with more respect and honesty”.

This positive relationship between education level and institutional trust is not found worldwide.While we do see such a connection in developed western democracies, it is less obvious in post-communist countries in Central and Eastern Europe and also in ‘new democracies’ in Latin America. We even find inverse relationships, where more highly educated persons have less trust than those with lower education levels [29, 30]. One possible explanation, offered by Van der Meer and Hakhverdian [31: 97], is that more highly educated persons are more critical of corruption than those with low education levels, so that the positive effect of education on trust is reduced or even inverted in countries where corruption is widespread. Also in the west, the relationship should not be taken as an absolute law, as shown by a cross-country comparative study that did not find a relationship between level of education and political trust in Germany, Switzerland and Spain. The researchers’ explanation of this finding is that ‘well-being’ was also included in the analysis as an explanatory factor, which may have blotted out the effect of education on trust [32: 112]. Despite these deviating outcomes, we assume here that respondents with a higher socioeconomic status have more institutional trust than those with a lower status.

Hypothesis 1: Socioeconomic status (SES) has a direct positive effect on institutional trust, whereby people with higher SES have more institutional trust.

### Perceived economic insecurity and institutional trust

Education, income, health and job market position are individual determinants of political and institutional trust. Social and economic factors also play a role, including especially the economic climate. It is generally thought that citizens trust the government while the economy is strong, and distrust the government when the economy deteriorates. If this is so, then we would expect that the COVID pandemic, with its massive economic impact on particular economic sectors, would also lead to less political and institutional trust.

The question is however whether people’s perception of the overall economic situation is more important than how they assess their own financial-economic situation. Wroe [33: 135] challenges the “…conventional wisdom (…) that citizens’ perceptions of the performance of the wider economy matter more than citizens’ perceptions of their own or their families’ financial situation”. Various studies show that it is not so much the assessment of the economic situation that determines the level of political trust, but much more the extent to which people feel vulnerable in case of an economic downturn [34]. Wroe [33] argues that a growing prosperity often goes hand in hand with growing economic insecurity for certain groups. An increasing economic insecurity would lead to a decline in political trust because citizens feel that the welfare state today offers too little protection against insecurities [33, 35]. Wroe [33] measures this economic insecurity by means of questions as to whether people are concerned about loss of job, reduced pension payments, inaccessibility of health care, and potential financial misery for their family. All these aspects of perceived economic insecurity correlate significantly with the level of political trust.

Further, positive assessments of the economy *and* the education level of respondents have a positive effect on political trust: the less concern about job loss and the higher the educational level, the higher the level of trust [33: 148]. Inspired by this research, we not only assume that economic insecurity – that is, the concern for loss of income due to the pandemic – has a negative effect on institutional trust, but also that this concern for loss of income occurs more strongly among lower status groups. These vulnerable groups more often work in sectors and occupations where homeworking is problematic, and where job and income security are lower [36]. People with lower incomes also have less financial reserves to rely on and the lower quality of housing is often a source of stress. That is why we expect that the economic insecurity as a result of the pandemic and the anti-corona measures fulfil a mediating role in the relationship between SES and trust. SES could therefore strengthen or weaken institutional trust through economic insecurity.

Hypothesis 2: Socioeconomic status (SES) has an indirect positive effect on trust through economic security, whereby people with a higher SES experience less economic insecurity and therefore have more trust.

### Discontent with government measures and institutional trust

Besides the effect of respondents’ socioeconomic status and their perceived economic insecurity due to the pandemic on institutional trust, their perceptions and assessments of government policies also – or perhaps particularly – affect their trust in the government. The premise here is that trust always pertains to a relationship between someone who trusts and someone who is trusted [37]. If the government pursue effective policies, is just and not corrupt, then political trust will be strong.Conversely, ineffective policies, injustice and especially corruption undermine political trust [38, 39]. Van der Meer and Dekker [39] add that it is not so much a matter of the factual government policies (as can be determined using objective criteria of effectiveness, justice or corruption) as of citizens’ *perceptions* regarding these aspects [40]. According to Van der Meer and Hackverdian [31: 82], this evaluative nature of political trust is ‘surprisingly understudied’ in the existing literature.

During 2020 and 2021, governments have taken unprecedented measures to counter and control COVID-19: from social distancing, avoiding contact, working at home as much as possible, avoiding public transport, shutting shops and hospitality outlets, to imposing a curfew (in many countries). Unsurprisingly, such drastic measures provoke discontent and resistance among citizens, particularly among those who are affected economically and/or feel overly constrained in their personal freedom. It is not without reason that riots occurred in several Dutch cities following the imposition of the curfew in January 2021. Following Van der Meer and Hackverdian [31], we presume that the positive or negative assessment of the corona policies pursued by public authorities correlates with people’s social status, particularly their level of education. According to both authors, education fulfils both a ‘norm-inducing’ and an ‘accuracy-inducing’ function [31: 86]. Higher educated persons are thought to be more critical of rule violations such as corruption on the one hand, and are better equipped to effectively process relevant information and to form well-considered judgements regarding corona policies on the other [41]. The first aspect would explain why corruption undermines political trust especially among higher educated citizens (and why higher educated people in corrupt countries in East Europe or Latin America have less trust). The second aspect could explain why shortcomings in government policies do not necessarily provoke frustration and discontent with the policies among the higher educated, and hence to less trust in the government and its implementing bodies. Instead, they might be more understanding of the difficult circumstances under which the policies are determined and implemented. Higher educated people furthermore tend to enjoy better resources (larger dwellings, more opportunity to work at home), so that they are better able to cope with the restrictions imposed by the corona policies.

In this study we not only assume that discontent with the corona policies has a negative effect on institutional trust, but also that discontent occurs more strongly among lower status groups. We therefore expect that discontent with the corona policies functions as mediator in the relationship between socioeconomic status and institutional trust. SES therefore strengthens or weakens institutional trust through discontent with the corona policies.

Hypothesis 3: Socioeconomic status (SES) has an indirect positive effect on trust via discontent with the corona policies, whereby people with a higher SES experience less discontent about the policies and hence have more trust.

#### Social status, economic insecurity, discontent with government measures and reduced trust

Finally, in this study we presume that all factors discussed so far are interrelated and reinforce each other. Respondents with a lower social status experience more economic insecurity, meaning that they are more concerned about possible income loss or have already suffered income loss.Respondents that experience more economic insecurity are also more negative about the corona policies pursued by the government [see 10]. The reason is, on the one hand, that they are more strongly affected by the economic consequences of government measures, for instance through loss of income, job or one’s own business, or on the other hand, because the corona measures hinder their efforts to improve their economic position, for instance by finding a job. We therefore presume that both economic insecurity and discontent with the corona policies act as mediators in the relationship between social status and institutional trust, simultaneously. Social status could hence both strengthen and weaken institutional trust through economic insecurity *and* through discontent with the corona policies.

Hypothesis 4: Socioeconomic status (SES) has an indirect positive effect on trust via economic insecurity and dissatisfaction with government policies, whereby people with a higher SES experience less economic insecurity, and therefore less dissatisfaction with the policies, resulting in stronger trust.

Based on the research findings and considerations described above, we present the conceptual research model below, where every letter represents a regression weight that indicates the relationship between the variables. More specifically, c represents the total effect of SES on trust and c’ represents the direct effect of SES on trust (controlled for economic insecurity and discontent with the corona policies). Accordingly, both weights are based on different linear regression analyses.

## Method and data

### Data, and sample and weighting

The data used in this study were obtained through a large-scale survey on the societal impact of COVID-19, performed by Kieskompas research agency. The data used here were collected between 28 October and 13 November 2020. This was the third measurement in a long-running study, with earlier measurements performed in April and July 2020, and a fourth measurement performed in March 2021 [see 10, 11]. The data were collected using Kieskompas’s permanent nationwide VIP panel. This panel is composed of a stratified random sampling of Dutch persons of voting age (18+). The survey was fielded among 48,329 members of the Kieskompas panel, and it was completed by 19,581 respondents (response rate of 40.5%). Additionally, three of the cities participating in the study (Amsterdam, The Hague and Rotterdam) performed additional activities to reach underrepresented groups by posting advertisements on Facebook and inviting specific groups with a weaker social status to participate. In Amsterdam, the questionnaire was also presented to the Amsterdam city panel. At the end of the field work period, the survey could also be completed using an anonymous participation link. In the end, the survey was completed by a sample of 22,696 respondents. To make the survey data representative for the (voting age) Dutch population, a weighting was performed retroactively. The goal of the weighting is to count under-represented groups in the database more often in order to more ‘accurately’ model the population and increase the representativeness of the sample. For example, the unweighted base contains more males (59%) than females (41%); in the weighted base, these sample frequencies have been updated to 50.2% males and 49.8% females. The weighting factor in our study varied from .06 to 16.35. By weighing the results in terms of gender, age, education region, ethnicity and voting behaviour, the data regarding these variables are (within the categories used) representative for the Dutch population aged 18 and over.

### Operationalisation of the variables

Institutional trust is the central dependent variable in this study. In the survey, respondents were asked to what level they have trust in national and local governments, *and* in important public health bodies such as the RIVM and GGD. Respondents could indicate whether they have (very) strong trust or (very) weak trust (1-5) in these four entities. The average of these items was taken as a measure of institutional trust. A Cronbach’s Alpha analysis shows a reliable scale (Cronbach’s Alpha .88).

Socioeconomic Status (SES) is operationalised as a combination score based on income and education level. The survey asked respondents both about their level of education (ranging from none to university level, Bachelor or Master), and about their net monthly income (ranging from less than €1150 for a single-person and €1600 for multiple-person households, to more than three times modal income (€5000 or more)). Using Principal Component Analysis (PCA), a factor score was created based on a least squares regression approach in order to reduce the variables to a single component. Note that according to the literature, education level plays a stronger role in explaining trust. Our data confirms this finding (we tested both education and income separately). However, income still explains a part of the variance in trust. Therefore we argue it is useful to use these variables in a combined measure. Further, in order to construct the SES variable, we have used the average income to impute the missing values for income.

#### Economic insecurity

Respondents were asked to what degree they felt anxious about losing their income due to the corona pandemic, using a 4-point scale where 1 represents ‘not anxious’, 2 ‘slightly anxious’, 3 ‘very anxious’, and 4 to indicate that the respondent had already lost (part of) his/her income due to the pandemic (and had therefore reached maximum economic insecurity). The rough scores of this variable were used to represent economic insecurity.

#### Discontent with government policies

Respondents were also asked for their opinion on the corona policies pursued by the Dutch government. They could indicate whether they (completely) agree or (completely) disagree (1 – 5) with respect to the following statements: “the Dutch government and media exaggerate the danger”, “the measures cause more damage than they prevent”, and “the government takes too little account of the economic and social consequences”. The answers to these three statements together form a reliable scale (Cronbach’s Alpha .83). The average of these items was taken as a measure of dissatisfaction with government policy. To be certain, we verified whether these items about dissatisfaction are not overly consistent with the items about the dependent variable of institutional trust. The four trust items and three dissatisfaction items cluster separately in a PCA with varimax rotation, and hence appear to be two different constructs.

#### Control variables

In all analyses reported below, controls are performed simultaneously for respondents’ age (in years) and gender. The descriptive information for all variables is presented in Table 1 below (see appendix).

**Table 1.** Descriptives of all variables in the analysis

|  | Share/<br>Mean | SD | Range | Missing<br>% |
| --- | --- | --- | --- | --- |
| Gender |  |  |  |  |
| Male | 49% |  |  | 0 |
| Female | 51% |  |  | 0 |
| Age | 50,2 | 17.54 | 19 - 96 | 0 |
| SES | 0 | 1 | -2.5 - 1.97 | 0 |
| Institutional trust | 3.36 | 1.01 | 1-5 | .7 |
| Economic insecurity | 1.55 | .82 | 1-4 | 1.0 |
| Discontent government policies | 2.64 | 1.10 | 1-5 | .5 |

**Note:** To construct the SES variable, we used the mean income to impute the missing values for income.

### Statistical analysis

The model shown in Figure 1 serves as the theoretical framework to test the associations between the different variables. In order to test this framework, we performed a serial mediation analysis using the PROCESS tool, a macro for SPSS [42], which incorporates the age and gender of the respondents as co-variates in the model. This procedure uses various ordinary-least-squares regression analyses to estimate the coefficients in the models and to determine the direct effects. Additional procedures were performed to estimate the indirect effects, specifically the multiplication of several regression weights. This approach uses bootstrapping (with replacement), with repeated sampling from the dataset (in our case, 5000 bootstrap-samples). In this way the tool generates an estimate for the sampling distribution of the indirect effects, in order to determine the reliability intervals of the estimated effects. The indirect effect is deemed to be statistically significant if the reliability interval does not contain a zero [cf. 42].

**Fig 1.**
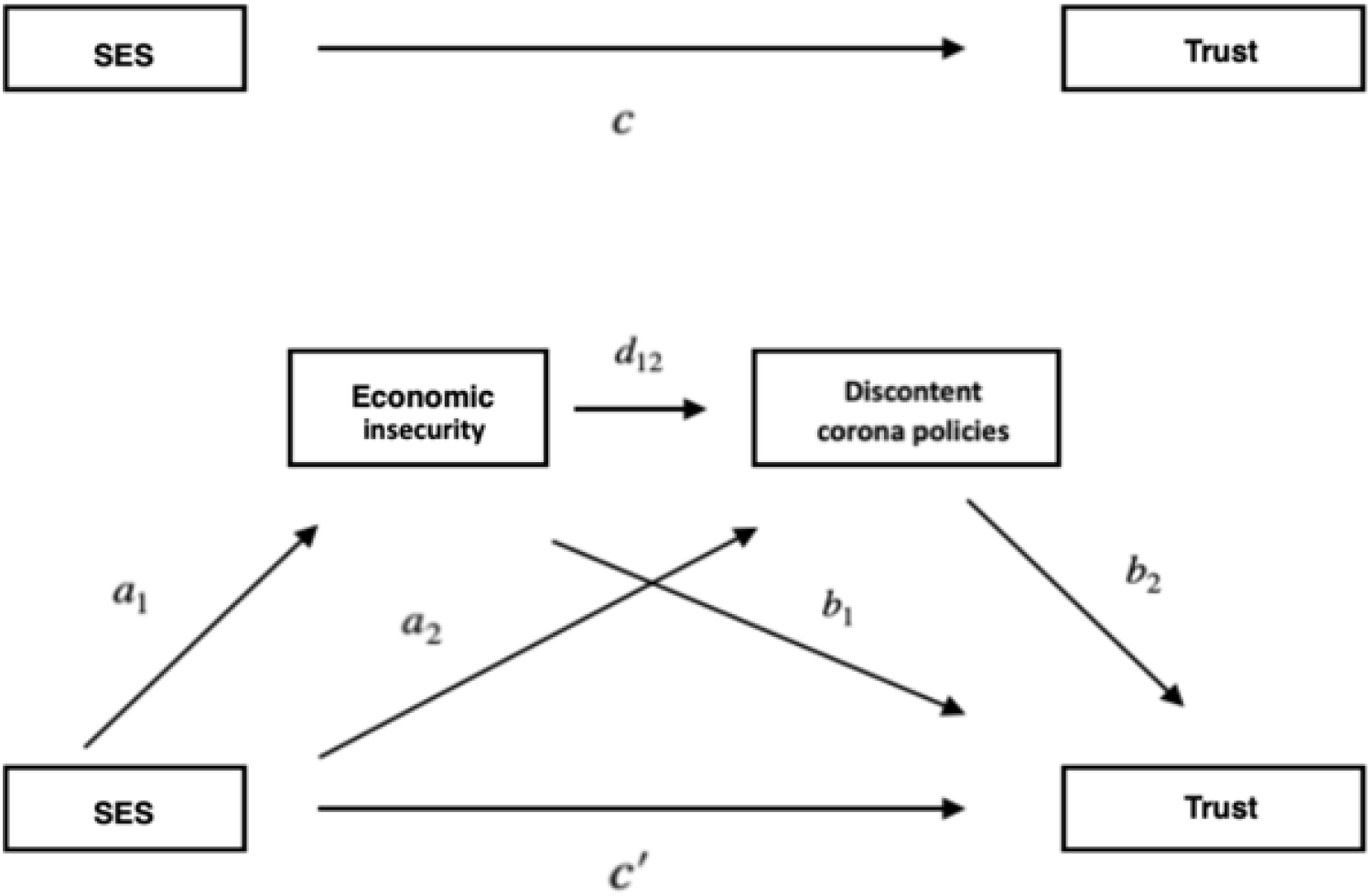
Conceptual research model

The analyses described below were performed on the unweighted survey data because the PROCESS tool precludes the use of weighted data. Although we make use of unweighted data in this article, the differences between the weighted and unweighted data are negligible. (Several regression analyses were performed manually on the weighted data to simulate the PROCESS results). Partly for this reason, and also given the size of the sample, we have confidence in the conclusions of our study.

## Results

### SES and trust

Our first research question is whether having a higher socioeconomic status (SES) in terms of education and income goes hand in hand with stronger trust in institutions (H1). The results indicate that, indeed, when SES increases, so does institutional trust (c = .22, p <.001). This result supports H1. Although this simple model explains 5.21% of the variance in trust, the explained variance increases to 30.97% when economic insecurity and discontent with the corona policies are included in the analysis. In this more elaborate model, SES still has a significant direct effect on trust, albeit slightly weaker (c’ = .135, p <.000). The reason is that part of the effect of SES now runs via the mediators (economic insecurity and discontent with the corona policies). See figure 2 for all standardized effects. For an overview of all un-standardized effects, see Table 2 in the appendix.

**Fig 2.**
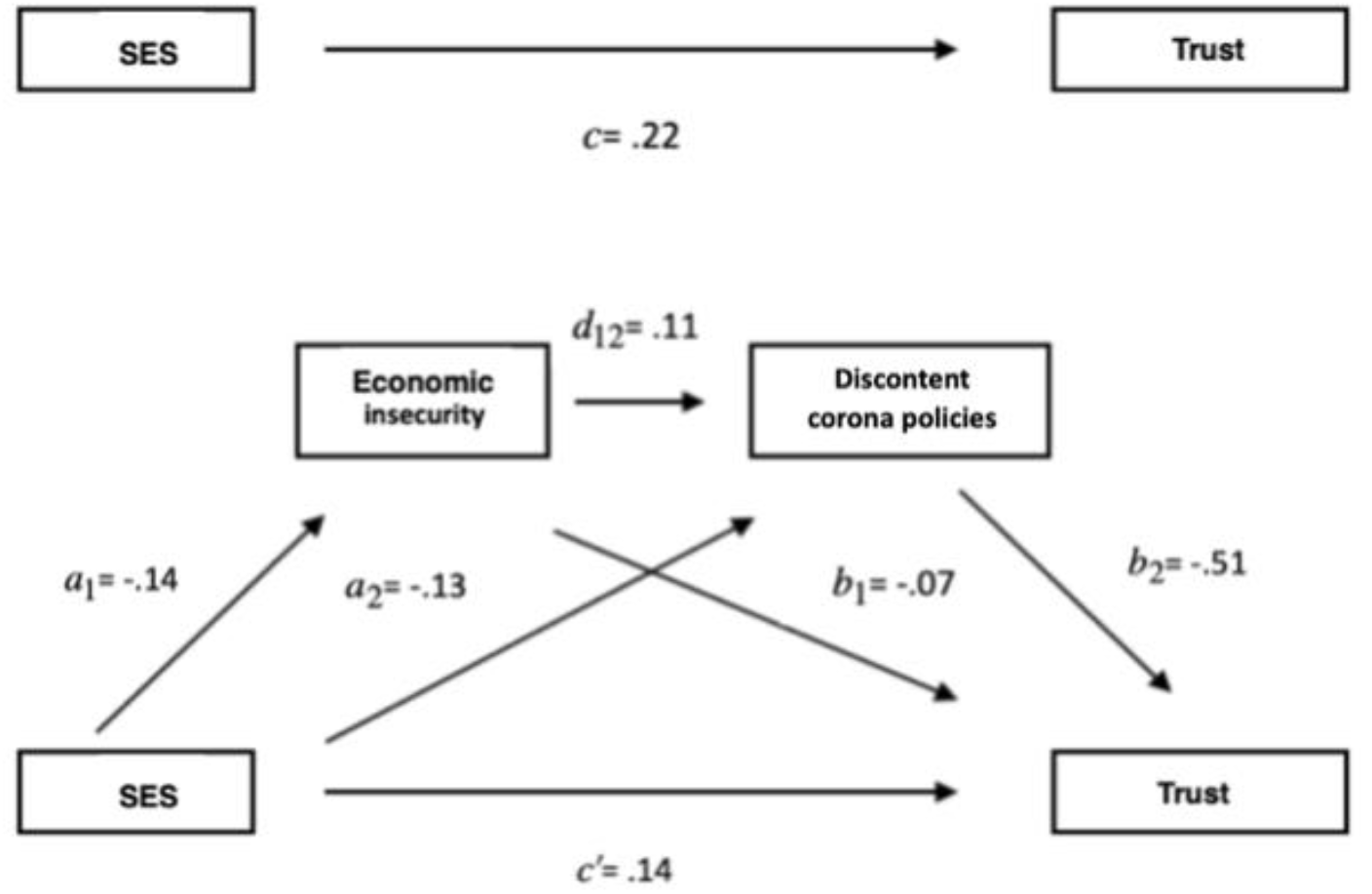
Standardised path coefficients (rounded off to two decimals)

**Table 2.** Unstandardised regression-coefficients and standard errors in various linear regression-analyses

| Predictor | Dependent variable |  |  |  |
| --- | --- | --- | --- | --- |
|  | Institutional trust | Economic insecurity | Discontent corona policies | Institutional trust |
|  | Coeff. (SE) | Coeff. (SE) | Coeff. (SE) | Coeff. (SE) |
| Constant | 3.598(.02) *** | 1.822(.02) *** | 2.507(.02) *** | 5.017(.02) *** |
| Age | -.003(.0004) *** | -.007(.0003) *** | -.008(.0004) *** | -.007(.0003) *** |
| Gender | -.009(.01) | .011(.01) | .108(.01) *** | .042(.01) *** |
| SES | **c** .215(.01) *** | **a<sub>1</sub>** -.118(.01) *** | **a<sub>2</sub>** -.141(.01) *** | **c'** .133(.01) *** |
| Economic insecurity | — | — | **d<sub>12</sub>** .142(.01) *** | **b<sub>1</sub>** -.081(.01) *** |
| Discontent corona policies | — | — | — | **b<sub>2</sub>** -.459(.01) *** |
| $R^2$ | .0521 | .0381 | .0506 | .3097 |
|  | F(3, 22396) = 410.31, p<.001 | F(3, 22396) = 295.67, p<.001 | F(4,22395) = 298.68, p<.001 | F(5, 22394) = 2009.55, p<.001 |

Note: All regression-coefficients rounded off to 3 decimals. p<.05\*, p<.01\*\*, p<.001\*\*\*.

### SES, trust and (perceived) economic insecurity

According to Hypothesis 2, the relationship between SES and trust is partly mediated by perceived economic insecurity. The results suggest that, as expected, respondents with higher SES experience less economic insecurity (*a*_1_= -.14, p <.000). Additionally, we see that respondents who experience more economic insecurity have less institutional trust (*b*_1_= -.07, p <.000). The results also show that the respondents’ socioeconomic status can strengthen (or weaken) institutional trust through economic insecurity, whereby a higher status strengthens trust via lesser economic insecurity (*a*_1_*b*_1_ = .0097, SE = .001, 95% CI = 0.008, .0117; For all direct and indirect effects, see Table 3 in the appendix). The results support H2; economic insecurity operates as a mediator in the relationship between SES and trust.

**Table 3.** Complete standardised estimates of the direct and indirect effects of SES on trust, with the associated reliability intervals.

| Total & Direct effect | Coeff. | Boot. SE | 95% | Figure-path |
| --- | --- | --- | --- | --- |
|  |  |  | reliability interval |  |
| Total effect of SES on Vert | .2190 | — | — | *c* |
| Direct effect of SES on Vert | .1352 | — | — | *c'* |
| **Indirecte effecten** |  |  |  |  |
| SES—>Ec_onz—>Vert | .0097 | .0010 | [.0079, .0117] | Ind<sub>1</sub>= (a<sub>1</sub>b<sub>1</sub>) |
| SES—>Onvr—>Vert | .0662 | .0035 | [.0591, .0733] | Ind<sub>2</sub>= (a<sub>2</sub>b<sub>2</sub>) |
| SES—>Ec_onz—>Onvr—>Vert | .0078 | .0006 | [.0066, .0091] | Ind<sub>3</sub>= (a<sub>1</sub>d<sub>12</sub>b<sub>2</sub>) |
| Total indirect effects | .0838 | .0037 | [.0765, .0912] | a<sub>1</sub>b<sub>1</sub> + a<sub>2</sub>b<sub>2</sub> + a<sub>1</sub>d<sub>12</sub>b<sub>2</sub> |

**Note:** The interval estimates are based on 5000 bootstrap samples.

### SES, trust and discontent

Hypothesis 3 holds that the association between socioeconomic status and trust is also mediated partly via respondents’ subjective evaluation of the corona policies. The results suggest that respondents with a lower SES are also less satisfied regarding the corona policies (*a*_2_ = -.13, p <.000). Additionally, we see that as discontent with the corona policies increases, institutional trust decreases (*b*_2_= -.51, p <.000). The results also show that SES can strengthen (or weaken) trust indirectly via discontent with the corona policies: a higher SES increases trust via less discontent with the corona policies (*a*_2_*b*_2_ = .0662, SE = .0035, 95% CI = .0591, .0733). These results support H3; discontent with the corona policies operates as a mediator in the relationship between SES and trust.

### Economic insecurity and discontent

Finally, our fourth hypothesis holds that perceived economic insecurity and discontent with the corona policies also influence the relationship between SES and trust in conjunction with each other (that is, simultaneously). The results show that economic insecurity has a positive association with discontent with the corona policies (*d*_12_ = .11, p <.001); respondents who experience more economic insecurity also experience more discontent with the corona policies. The indirect effect of SES on trust via economic insecurity and discontent with the corona policies is also significant (*a*_1_*d*_12_*b*_2_ = .0078, SE = .0006, 95% CI = .0765, .0912). These results support our fourth hypothesis that there is an indirect and positive association between socioeconomic status and institutional trust via economic insecurity and discontent with the corona policies, whereby a higher SES is associated with less economic insecurity, and hence with less dissatisfaction, resulting in stronger institutional trust.

Although all our hypotheses are supported by the results, some effects are stronger than others, as shown in Figure 2. For instance, discontent with the corona policies shows the strongest association with institutional trust. Table 2 (in the appendix) furthermore shows that the explained variance of economic uncertainty (3.81%) and discontent with the corona policies (5.06%) is lower compared to institutional trust (30.97%). This implies that SES and economic uncertainty – despite statistical significance – do not explain the largest share of the variance in the associated variables. It does seem clear nevertheless that (a) SES has an effect on economic insecurity, (b) discontent with the corona policies is still affected by both SES and perceived economic insecurity, and (c) the effect of these variables plays a non-negligible role in the explanation of the level of institutional trust, both directly and indirectly (through dissatisfaction).

### The role of age and gender

All analyses were controlled for age and gender simultaneously. As Table 2 shows (see appendix), in some cases there is a significant association between these control variables and the other variables in this study. For instance, elderly respondents experience less economic insecurity (β= -.01, p <.001) and also feel less discontent with the corona policies (β= -.01, p <.001). Nevertheless, there is a negative association between age and institutional trust. Although elderly respondents experience less economic insecurity *and* are less dissatisfied with the corona policies, they still have less institutional trust than younger respondents (in both the simplified and the elaborate model). The (negative) effect of age on trust becomes stronger as we keep the degree of economic insecurity and of dissatisfaction constant for the respondents in the elaborate model (β= -.007, p <.001). Hence, higher age in alle cases corresponds with less trust, and this effect becomes stronger when we keep the degree of economic insecurity and dissatisfaction constant for the respondents.

We do not find any statistically significant differences between males and females regarding the degree of perceived economic insecurity (β= .01, p = .29). Males do show more discontent with the corona policies than females (β= .11, p <.001), but notwithstanding this difference, males and females show the same level of institutional trust. It is only when we keep the degree of dissatisfaction and the degree of economic insecurity constant for both sexes, that males appear to have more trust than females (β= .04, p <.001, Table 2). We can therefore conclude that the differences between the sexes are minimal in this study.

## Conclusion and discussion

The main research question of this study was whether the usual research outcomes regarding political and institutional trust, namely that the higher educated and higher income groups (the societal ‘winners’) show more trust, also applies during the corona pandemic. We focused on institutional trust, that is trust in the government (both national and local) and in public health organizations (GGD and RIVM). However, we not only investigated the relationship between respondents’ social status and the level of institutional trust, but also the role of perceived economic insecurity and the respondents’ subjective evaluation of the corona policies pursued by the Dutch government. For our study we used survey data collected in November 2020, when institutional trust in the Netherlands had weakened considerably.

The main outcome of our study is that SES has both a direct and indirect effect on the level of institutional trust. In line with earlier research, SES was found to have a direct effect on institutional trust. Also when respondents experience the same degree of economic insecurity and discontent with the policies, those with a higher SES show stronger institutional trust. One possible explanation is that people in higher social positions tend to be more optimistic and positive about public authorities, while people in more vulnerable social positions are more cynical, pessimistic and distrustful. It has also been noted that people in higher social positions generally experience less insecurity and problems in their life and therefore feel better protected by public authorities than less successful people.

This direct effect is strengthened by the indirect relationship between SES and both other factors. We concluded that SES is also associated with both perceived economic insecurity and discontent with the corona policies, which in turn predicted the level of institutional trust. People with higher SES experience less economic insecurity and are less dissatisfied regarding the corona policies, and accordingly show more institutional trust. The corona pandemic clearly reveals that people with lower education levels and lower incomes are hit hardest by the economic consequences of corona, for instance through loss of job or income *or* because they fear such loss. People with low education levels and flexible jobs, or self-employed people with comparable positions, often lost their jobs (unless they worked in the parcels or meals delivery sector). Higher educated people, on the other hand, are generally much better positioned to work at home and could hence continue to live and work relatively ‘normally’. Their permanent jobs furthermore provided greater income security. We also found that the perceived economic insecurity is related to discontent with the corona policies. People who experience a stronger degree of economic insecurity tend to be more negative about the policies pursued by the government. These are all factors that contribute to a weaker level of institutional trust among lower SES groups.

Additionally, we found that a more advanced age is also associated with less trust, even though older respondents on average experience less economic insecurity and less discontent with the corona policies. Younger people (in our study, respondents aged 18 to 34) on average show more trust in institutions, although they are hit harder by both the economic and mental effects of the pandemic than older respondents [43]. Apparently, younger Dutch nationals have more trust in the ability of government and public health bodies such as the RIVM and GGD to adequately manage the pandemic than elderly compatriots. Finally, we found little difference in institutional trust between males and females, although the latter do show more discontent with the corona policies. Only if the degree of economic insecurity and dissatisfaction with the policies were the same for both sexes, then males would appear to have *more* institutional trust than females. This finding is striking, since available research shows that females generally have slightly stronger political or institutional trust than males [44]. Perhaps the “triple burden” experienced by many women during the pandemic (looking after work, the household, *and* having the children at home) can explain this deviating outcome.

The relevance of this study and our findings is two-fold. The scientific relevance is that we present a more complete theoretical model that allows us to better explain differences in institutional trust, compared to only considering respondents’ social status (their level of education and/or income) as a predictor of trust. The two theoretically substantiated additions to this classic relationship – perceived economic insecurity [33], and especially the subjective evaluation of governmental policies [31] – considerably increase the model’s explanatory power. The model using only SES (and both control variables) as predictor explained 5.21% of the variance in trust, while this increased to 30.97% after including perceived economic insecurity and discontent with the corona policies in the analysis. Particularly the latter factor is strongly associated with institutional trust. SES continues to explain part of the variance in trust, but this relationship is reinforced through perceived economic insecurity and especially discontent with the corona policies. Both SES and economic insecurity have both a *direct* and *indirect* role in explaining the level of institutional trust, and more strongly so than if we were to only consider the individual (direct) effects on trust of the two factors. In sum: because all the relevant variables are interconnected, we may conclude that the complete picture as presented in Figure 1 offers a better and more comprehensive picture of the hypothetical relationships.

The societal relevance of our study is that, particularly in this time of the corona pandemic, institutional trust is crucially important. Governments are imposing drastic social and economic measures to combat the virus. The extent to which people are willing to obey the rules, and to which they are prepared to be vaccinated, is closely associated with their trust in government and in important public health bodies such as the RIVM and GGD [cf. 10, 45]. It is therefore very important that the government also gains (or regains) the trust of lower social status groups, of people who have personally suffered the economic consequences of the pandemic, *and* of those who are critical of the government’s corona policies. This implies on the one hand, a policy aimed at strengthening the economic security of vulnerable groups, and on the other an adequate public health service and clear communication policy to explain to groups why certain measures are vital. As regards communication policy, the fundamental and practical guidelines formulated by Tiemeijer [46] are relevant, including fundamental principles such as *legitimacy* (‘be truthful, just and reasonable’) and *effectiveness* (‘be clear and unambiguous’) and practical matters such as ‘know your target group’ and ‘offer hope, small intermediate steps, celebrate achievements, acknowledge emotions’.

### Limitations of the study

This study, like every study, has certain limitations. First, the cross-sectional research design limits the extent to which cause-effect relationships can be deduced from the findings. For example, we describe the relationship between discontent with the policies and institutional trust, but cannot pinpoint the exact causal relationships. Satisfaction regarding the policies can lead to more institutional trust because people take a positive view of the measures. Yet one could also argue that people who have more institutional trust will naturally be inclined to take a positive view of governmental policies. In other words, the causal direction of the observed relationships is not immediately apparent. Still, our research model as presented in Figure 1 is theoretically grounded in the literature, and we discuss the outcomes of the model under the assumption that these are the correct causal directions.

Another limitation pertains to the operationalisation of some of the variables. Thus, the variable ‘income’ lacked several values (12.7%). To construct the SES variable, we used averages to impute these missing values, and such estimates could of course be wide of the mark. However, we did not want to drop respondents from the analysis because it would also rule out using other information about these respondents (regarding economic insecurity, discontent with the corona policies and institutional trust) in our analysis.

Finally, we limited the scope of this study to three factors to explain the differences in trust. There are however other factors that could also contribute to explaining differences in trust, such as political orientation and psychological factors (e.g. stress) [10]. Our recommendation for further research is accordingly to include such factors as covariates in future analyses. Further, the outcome measurement in our study pertains to the level of ‘trust’. It would be interesting for future studies to also include the level of ‘distrust’ as an outcome measurement, in order to test to what extent the results do or do not overlap.

## Data Availability

The data underlying the results presented in the study are available from the
corresponding author.

## Appendix

